# SARS-CoV-2 infection and COVID-19 severity in individuals with prior seasonal coronavirus infection

**DOI:** 10.1101/2020.12.04.20243741

**Authors:** Saurabh Gombar, Timothy Bergquist, Vikas Pejaver, Noah E. Hammarlund, Kanagavel Murugesan, Sean Mooney, Nigam Shah, Benjamin A. Pinsky, Niaz Banaei

**Author notes:** **Corresponding Author:** Niaz Banaei, MD, 3375 Hillview Ave, Rm. 1602, Palo Alto, Ca 94304.

## Abstract

A sizable fraction of healthy blood donors have cross-reactive T cells to SARS-CoV-2 peptides due to prior infection with seasonal coronavirus. Understanding the role of cross-reactive T cells in immunity to SARS-CoV-2 has implications for managing the COVID-19 pandemic. We show that individuals with documented history of seasonal coronavirus have a similar SARS-CoV-2 infection rate and COVID-19 severity as those with no prior history of seasonal coronavirus. Our findings suggest prior infection with seasonal coronavirus does not provide immunity to subsequent infection with SARS-CoV-2.

---

Understanding correlates of protective immunity against SARS-CoV-2 remains a top priority for management of the COVID-19 pandemic. Recent studies have identified a durable cross-reactive memory T cell immune response to SARS-CoV-2 in healthy blood donors, including those who donated prior to COVID-19 pandemic, which react with comparable affinity to seasonal coronaviruses (OC43, HKU1, 229E and NL63) *(1-3)*. These cross-reactive T cells are detected in 20 to 50% of unexposed blood donors from geographically diverse locations such as Germany, Netherlands, Singapore, USA, and UK, and are thought to represent a lasting cellular immune response to past seasonal coronavirus infection *(1, 2, 4-6)*. The significance of a pre-existing cross-reactive cellular immune response to SARS-CoV-2 in terms of protection against infection with SARS-CoV-2 and severity of COVID-19 in those that do get infected remains speculative but has great implications for managing the pandemic and a rational vaccine strategy for the global community.

To investigate the impact of past infection with seasonal coronavirus on subsequent susceptibility to SARS-CoV-2 and severity of COVID-19, we retrospectively searched the electronic health records of 2.9 and 5.4 million patients at Stanford Health Care and University of Washington, respectively, from January 1, 2016 to October 15, 2020 to identify individuals who had both seasonal coronavirus RT-PCR result prior to February 1, 2020 (pre COVID-19 pandemic), and subsequent SARS-CoV-2 RT-PCR result after February 1, 2020 (COVID-19 pandemic). At least one positive result, and the last negative or positive result in those with multiple seasonal coronavirus RT-PCR results was included. In total 2768 individuals (Northern California, n=1419; Washington, n = 1281) met the inclusion criteria.

In 302 patients with a documented history of positive seasonal coronavirus test results, 3.3% (10) subsequently tested positive for SARS-CoV-2 compared with 3.1% (77) in 2466 patients with a history of negative seasonal coronavirus result (P = 0.86, Chi-Square test) (Table 1). Similar results were obtained when seasonal coronavirus results were analyzed per year to determine if more recent infection had an impact on SARS-CoV-2 positivity rate (Supplementary Table). We also did not observe a higher SARS-CoV-2 positivity rate in patients ≥65 vs <65 years old in groups with a history of either positive or negative seasonal coronavirus result (data not shown). Lastly, in those with COVID-19, the proportion of patients with severe (defined as hospitalized and intubated) or moderate (defined as hospitalized but not intubated) infection was comparable to those with history of positive and negative seasonal coronavirus test results (40.0% (4/10) vs 48.1% (37/77), P = 0.63) (Table 2).

**Table 1.** SARS-CoV-2 RT-PCR positivity rate in patients with history of seasonal coronavirus prior to COVID-19 pandemic.

|  |  |  | Post-Pandemic SARS-CoV-2 |  |  |
| --- | --- | --- | --- | --- | --- |
|  |  |  | Positive | Negative | P Value |
| Pre-Pandemic Seasonal CoV | Northern California | Positive | 4.3% (7) | 95.7% (157) | 0.81 |
|  |  | Negative | 3.9% (49) | 96.1% (1211) |  |
|  | Washington | Positive | 2.2% (3) | 97.8% (135) | 0.95 |
|  |  | Negative | 2.3% (28) | 97.7% (1178) |  |
|  | Combined | Positive | 3.3% (10) | 96.7% (392) | 0.77 |
|  |  | Negative | 3.1% (77) | 96.9% (2389) |  |

**Table 2.** Severity of COVID-19 in patients with history of seasonal coronavirus prior to COVID-19 pandemic.

|  |  |  | Post-Pandemic SARS-CoV-2 |  |  |
| --- | --- | --- | --- | --- | --- |
|  |  |  | Mild | Moderate/ Severe | P Value |
| Pre-Pandemic Seasonal CoV | Northern California | Positive | 71.4% (5) | 28.6% (2) | 0.83 |
|  |  | Negative | 67.3% (33) | 32.7% (16) |  |
|  | Washington | Positive | 33.3% (1) | 66.7% (2) | 0.75 |
|  |  | Negative | 25.0% (7) | 75.0% (21) |  |
|  | Combined | Positive | 60.0% (6) | 40.0% (4) | 0.63 |
|  |  | Negative | 51.9% (40) | 48.1% (37) |  |

Our findings suggest past infection with seasonal coronavirus, and thus by inference cross-reactive memory T cell immune response to SARS-CoV-2, do not provide protective immunity against subsequent infection with SARS-CoV-2 infection and also do not modulate the severity of COVID-19 in those that get infected by SARS-CoV-2. Our findings are consistent with the lack of protection against SARS-CoV-2 infection in children with prior infection with seasonal coronavirus based on seropositivity *(7)*. Retrospective studies with larger number of participants and prospective studies in individuals with pre-existing cross-reactive memory T cells are needed to confirm lack of protective immunity to SARS-CoV-2 or modulation of COVID-19 severity in individuals with pre-existing cross-reactive T cells to seasonal coronaviruses.

## Data Availability

The data used to generate these findings can be accessed from Stanford University or University of Washington subsequent to obtaining a data usage agreement (DUA).

