## Supplementary Table for "SARS-CoV-2 infection and COVID-19 severity in individuals with prior seasonal coronavirus infection"

**Supplementary Table.** SARS-CoV2 RT-PCR positivity rate in patients with history of seasonal coronavirus testing prior to COVID-19 pandemic.

| | | | | Post-Pandemic SARS-CoV-2 | | |
|---|---|---|---|---|---|---|
| | | | | **Positive** | **Negative** | ***P* Value** |
| **Pre-Pandemic Seasonal CoV** | **2016** | **Northern California** | **Positive** | 0 | 0 | NA |
| | | | **Negative** | 0 | 0 | |
| | | **Washington** | **Positive** | 0% (0) | 100% (10) | NA |
| | | | **Negative** | 3.6% (2) | 96.4% (55) | |
| | | **Combined** | **Positive** | 0 | 100% (10) | NA |
| | | | **Negative** | 3.6% (2) | 96.4% (55) | |
| | **2017** | **Northern California** | **Positive** | 7.1% (1) | 92.9% (13) | 0.78 |
| | | | **Negative** | 5.3% (7) | 94.7% (124) | |
| | | **Washington** | **Positive** | 0 | 100% (32) | NA |
| | | | **Negative** | 3.3% (5) | 96.7% (147) | |
| | | **Combined** | **Positive** | 2.2% (1) | 97.8% (45) | 0.5 |
| | | | **Negative** | 4.2% (12) | 95.8% (271) | |
| | **2018** | **Northern California** | **Positive** | 5.2% (3) | 94.8% (55) | 0.44 |
| | | | **Negative** | 3.2% (12) | 96.8% (365) | |
| | | **Washington** | **Positive** | 3.6% (1) | 96.4% (27) | 0.65 |
| | | | **Negative** | 2.2% (6) | 97.8% (266) | |
| | | **Combined** | **Positive** | 4.7% (4) | 95.3% (82) | 0.34 |
| | | | **Negative** | 2.8% (18) | 97.2% (631) | |
| | **2019** | **Northern California** | **Positive** | 3.3% (3) | 96.7% (89) | 0.81 |
| | | | **Negative** | 4.0 % (30) | 96.0% (722) | |
| | | **Washington** | **Positive** | 2.9% (2) | 97.1% (66) | 0.91 |
| | | | **Negative** | 2.1% (15) | 97.9% (710) | |
| | | **Combined** | **Positive** | 3.1% (5) | 96.9% (155) | 0.96 |
| | | | **Negative** | 3.0 % (45) | 97.0% (1432) | |
| | **2016-2019** | **Northern California** | **Positive** | 4.3% (7) | 95.7% (157) | 0.81 |
| | | | **Negative** | 3.9% (49) | 96.1% (1211) | |
| | | **Washington** | **Positive** | 2.2% (3) | 97.8% (135) | 0.95 |
| | | | **Negative** | 2.3% (28) | 97.7% (1178) | |
| | | **Combined** | **Positive** | 3.3% (10) | 96.7% (392) | 0.77 |
| | | | **Negative** | 3.1% (77) | 96.9% (2389) | |

2019 included data from January 2020
